# Determinants of Eye Care Service Utilization Among Malawian adults; A Secondary Analysis of the Malawi fifth Integrated Household Survey 2019-2020

**DOI:** 10.1101/2022.11.09.22282125

**Authors:** Thokozani Mzumara

**Affiliations:** Unicaf university, Zambia; Mzimba North District Hospital, Malawi; Mzuzu university, Malawi

**Keywords:** access to health, utilization, eye care use, health seeking behavior, sociodemographic determinants

## Abstract

**Purpose:** The aim of the study was to assess the self-reported Eye Care Utilization (ECU) and associated demographic factors among Malawians adults.

**Methods:** This study utilized secondary data from the Malawi Integrated Health Survey (IHS) 5 2019-2020. We entered age, sex, level of education, residency(urban/rural) and having chronic disease into a logistic regression model and used a confusion matrix to predict the accuracy of the model.

**Results:** The prevalence of ECU was 60.6% (95% CI [60.0,61.2]). The model was statistically significant and correctly classified 72 % of the cases. A logistic regression showed that ECU was positively associated with education compared to none (OR=3.6, 95 % CI [3.104-4.097], p<0.001, (OR =6.6), CI [5.927,7.366], p<0.001), male gender compared to females (OR= 1.2, 95% CI [1.104,1.290], p<0.001), urban residence compared to rural areas (OR= 1.2, 95% CI [1.118,1.375], p<0.001). But It was negatively associated with age compared to young adults, middle age (OR =7.5, 95% CI [6.782,8.476], p<0.001), older age (OR=0.9, 95 % CI [0.866,1.035], p<0.001), and having chronic diseases (OR=0.6, 95 % CI= [0.547,0.708], p<0.001).

**Conclusion:** Social support, women empowerment, education, and mobile clinics are key strategic areas that would increase Access to eye care in Malawi. Further studies can investigate ECU among the pediatric population.

## INTRODUCTION

In 2020, 596 million people had visual impairment worldwide and 90 % lived in low income countries. ^[1]^ Approximately, 26 million people are visually impaired and 3 million blind in sub Saharan region. ^[2]^ In Malawi, 3.3 % of the population is blind compared to 1.01 % in America.^[3,4]^

Accordingly, visual impairment is directly related to access, Utilization and poverty. ^[2]^ Specifically, Eye care Utilization (ECU) refers to any eye activity for the purpose of treatment, prevention and health promotion.^[5]^ Sociocultural determinants such as norms, autonomy and decision making affect the utilization of healthcare.^[6]^ Hence, healthcare solutions heavily rely on human behavior.^[7]^In particular, Health Seeking Behavior (HSB) is driven by accessibility to care, perceived severity and susceptibility and perceived benefits of treatments.^[8]^ The model distinguishes two dimensions: potential and realized access.^[9]^ Furthermore, The Andersons model of healthcare classifies utilization into three dynamic forces: the predisposing, need, enabling factors.^[10]^ In summary, decision making with respect to healthcare mirrors social norms.^[7]^

Research suggests that low uptake of services is a major hindrance to the attainment of universal health. ^[11]^ In developing countries uptake of eye care service remains a challenge. ^[5,12]^ Globally, ECU varies widely ranging from 18% to 83%.^[5,13,14]^ Besides, studies have found discrepancies within countries with respect to urban and rural residents, sex, comorbid, age and education.^[9]^

Despite free eye care services in Malawi,^[15]^ health service uptake is lower ^[16]^. Evidently, government assistance does not guarantee utilization of services among poor individuals. ^[17]^ For instance, 40 % of Malawians initiate self-treatment for eye problems. ^[18]^ In contrast, eye problem is strongly associated with HSB. ^[19]^ However, the pattern of ECU in Malawi remains a conundrum.

Accordingly, utilization reflects coverage of health service and is a vivid indicator of health system performance. ^[20]^ Again, proper use of eye care services is key to dismantling the burden of visual impairment in developing countries. ^[5]^ Therefore, this paper aims at investigating the pattern of ECU and its associated demographic factors among Malawians. The study findings can help address systematic exclusion of vulnerable groups by shaping health delivery programs to ensure equal access to health.

## Methods and materials

This was a desk based review of the Malawi fifth Integrated Household Survey (IHS) 2019-2020.^[21]^ The survey was implemented between April 2019 to April 2020.

### Sampling procedure

The survey sampling frame, based on the 2018 Malawi Population and Housing Census,^[21]^ The survey utilized a stratified two stage sampling technique and interviewed 28388 Adults aged 15 years and above.

The sample excluded populations in institutions such as prisons, hospitals and military barracks.

### Sampling weights

To obtain a population representation of the sample estimates from the IHS5, we multiplied the data by sampling weights to analyze means, proportions and ratios.. ^[21]^

### Data management

This review extracted age in years, sex(male/female), residence(rural/urban), highest level of education, and sampling weights. Next, age of participants was re-grouped into three sets comprising of young adults (15-34 years), middle aged (35-59 years), and older adults (60 years and above). Next, we recoded the level of Education into five categories namely none (no education and don’t know), primary (primary school leaving certificate PSLCE), secondary (including A level), tertiary, (diploma, first degree, masters, phD). Furthermore, the paper extracted information related to symptoms experienced for the previous 2 weeks (yes/No), name of symptom, the action taken to relieve the symptom and presence of chronic illness.

To determine ECU we recoded the action taken to relieve the symptom as 1 if the individual sought care at a government or private facility including church/mission facility, village clinic and pharmacy store. All other options including self-care and use of stock medicine and other non-orthodox practices were recoded as 0.

### Analysis strategy

The data was entered in Statistical Package for Social Sciences (SPSS) version 26 (SPSS Inc., Chicago, Illinois). We executed descriptive statistics including mean and standard deviation, frequency and proportion. We illustrated the data graphically using tables and figures. Proportional data was analyzed using chi-square and we considered the value of p less than 0.05 significant. To estimate the probability ECU occurring, we entered the variables into binary logistic regression model employing entry method. The probability cut off was set at 0.5%, as such probabilities greater than 0.5 was classified as ECU (event occurring) else no ECU (event not occurring). To assess the prediction classification model, the study employed the confusion matrix technique and calculated the predictive values accuracy, specificity and sensitivity of the model. Confusion matrix is a machine learning technique. ^[22]^

### Ethics

The study did not require institutional review because it used de-identified publically available data from the world bank accessible through https://microdata.worldbank.org/index.php/catalog/2939.

## Results

### Characteristics of study participants

The participants age ranged from 15 to 118 years and the mean age was 34.36 (SD=16.973). The most frequent age group was young adults 59% (5,955,521) while older adults comprised 10% (1,024,206) only. According to gender, there were 4,724,880 (47 %) males and 5,363,948 (53 %) females. About 5,660,836 (56 %) of the adults were married whereas 621,557 (6%) were widows/widowers. Majority 8,375,415 (83%) were from the rural areas and the remainder 1,713,412 (17%) from the urban areas. According to region, 1,371,898 (13 %) were from the North, and 4,366,155 (43 %) from the South. Concerning the highest education qualification obtained, 160,710 (1.6%) seized tertiary education compared to 5,795,168 (57%) who attained PSLC.

### 8.2 Prevalence of eye diseases 2 weeks preceding the survey

In general, 2,734,768 (27 %) participants reported having experienced an illness or injury 2 weeks earlier to the study. Out of these, 27,336 (0.3% of 2,734,768) complained of ocular symptoms. The mean age of participants with ocular symptoms was 47.68 (SD= 24.39). The prevalence of eye symptoms was lowest among young adults 11,118 (0.8%%) and highest among older adults 11,155 (2.4% of 459593) (p<0.000). A larger population of females 19323 (1.2% of 1,576,744) compared to males 8,014 (0.7% of 1,158,024) (p< 0.001) had ocular symptoms. According to region, the prevalence was higher in the South 12,378 (1 % of 1,238,222) and lowest in the North 3084 (0.9 % of 334,402). A total of 4135 (1.2% of 344,954) were from the urban area, while 23,202 (1% of 2,389,814) were from the rural areas. (p<0.001) Table 1.

**Table 1.**
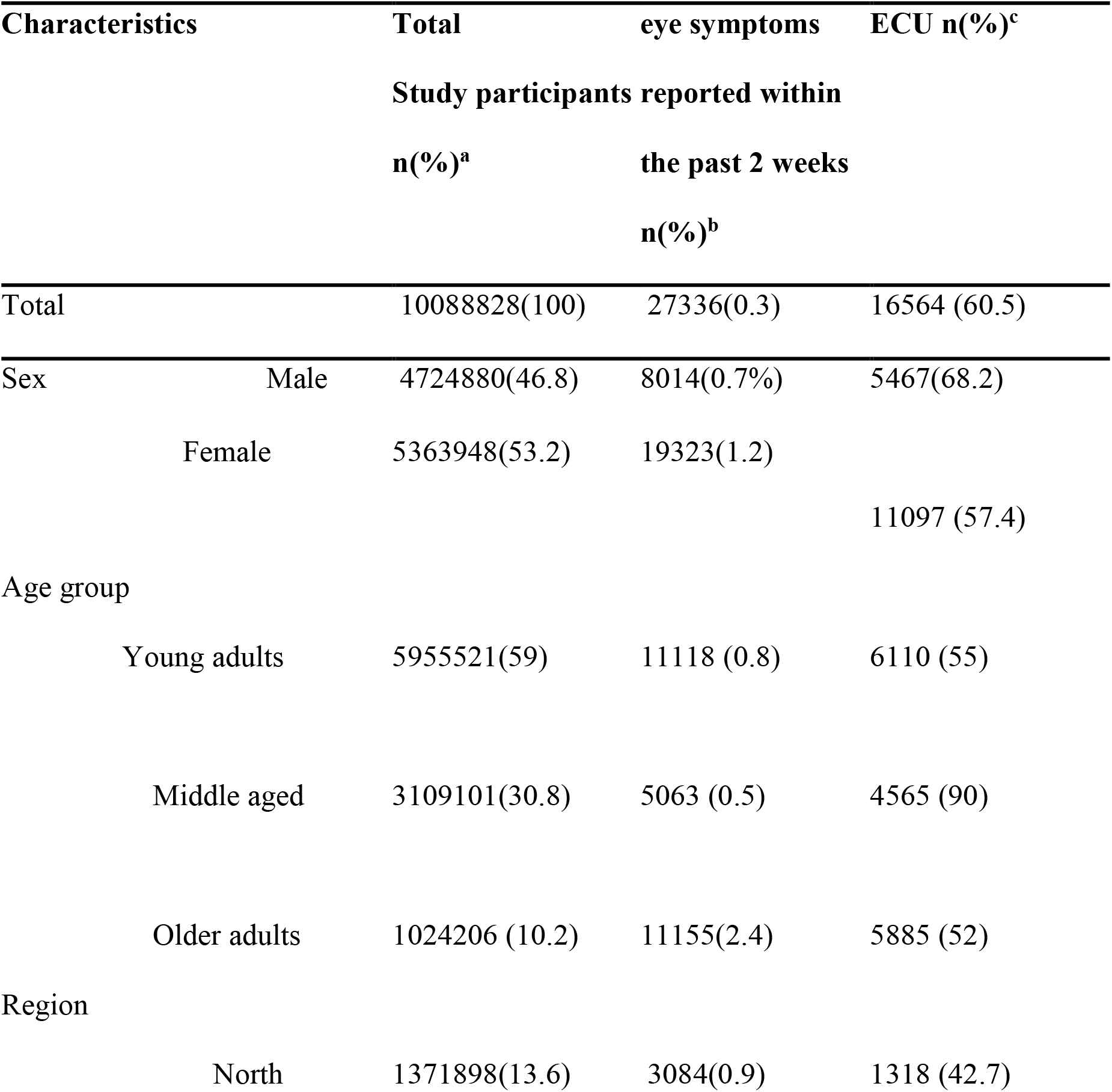
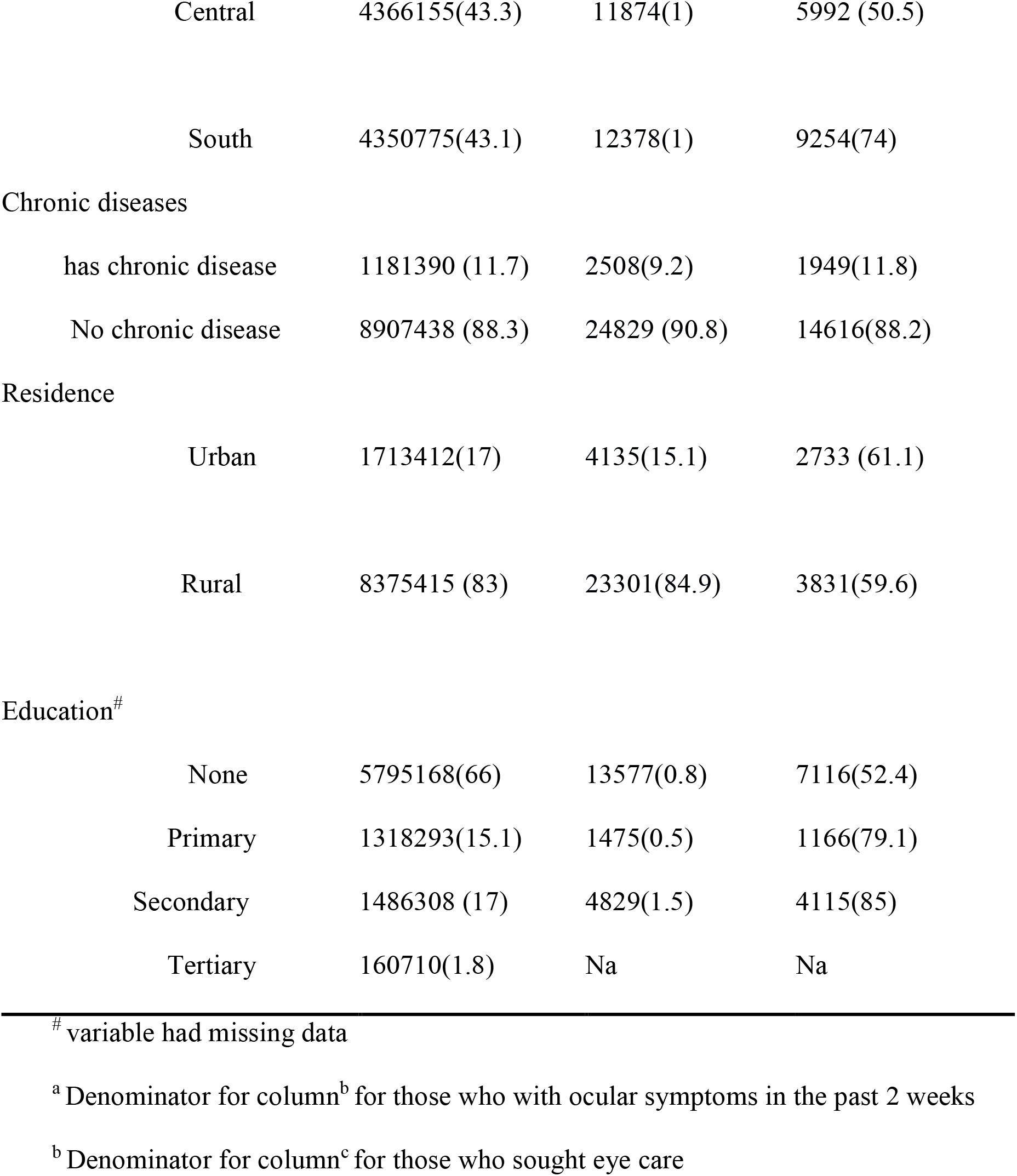
characteristics of study participants with eye symptoms and participant’s ECU.

### Places where participants sought help

Table 2 below portrays the distribution of individuals with eye problems according to where they sought help. A total of 16,564 (60.6 % of 27,336) (95% CI [60.0,61.2]) sought eye care from a medical/health facility. Out of 16,564 that sought care from a health facility, 14,173 (85%) was at a government facility where as 463 (2.7%) obtained drugs from their local pharmacy. Among those who did not seek care, 2,950 (46%) attributed it to lack of funds while 3,393 (54%) did not think it was a serious illness.

**Table 2.**
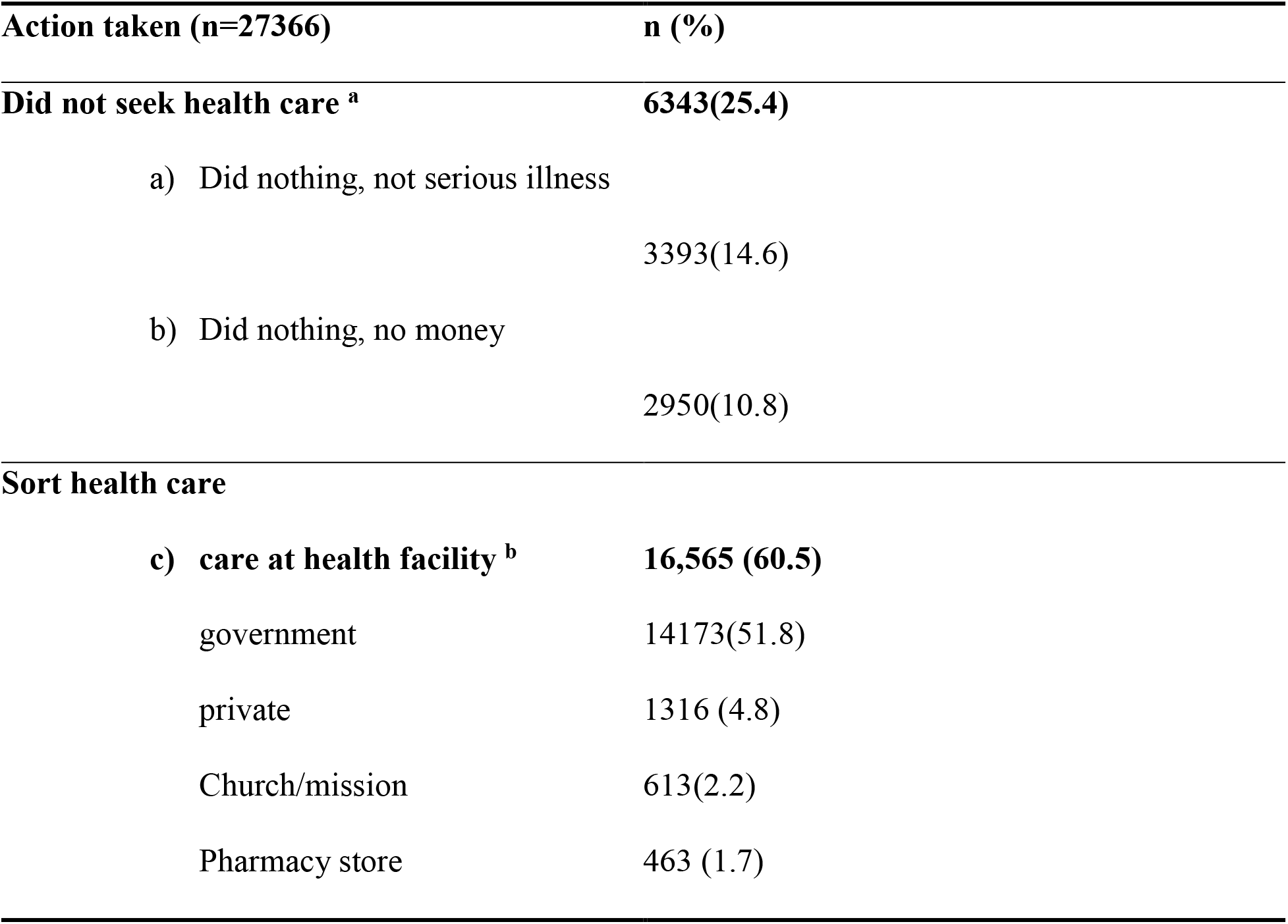
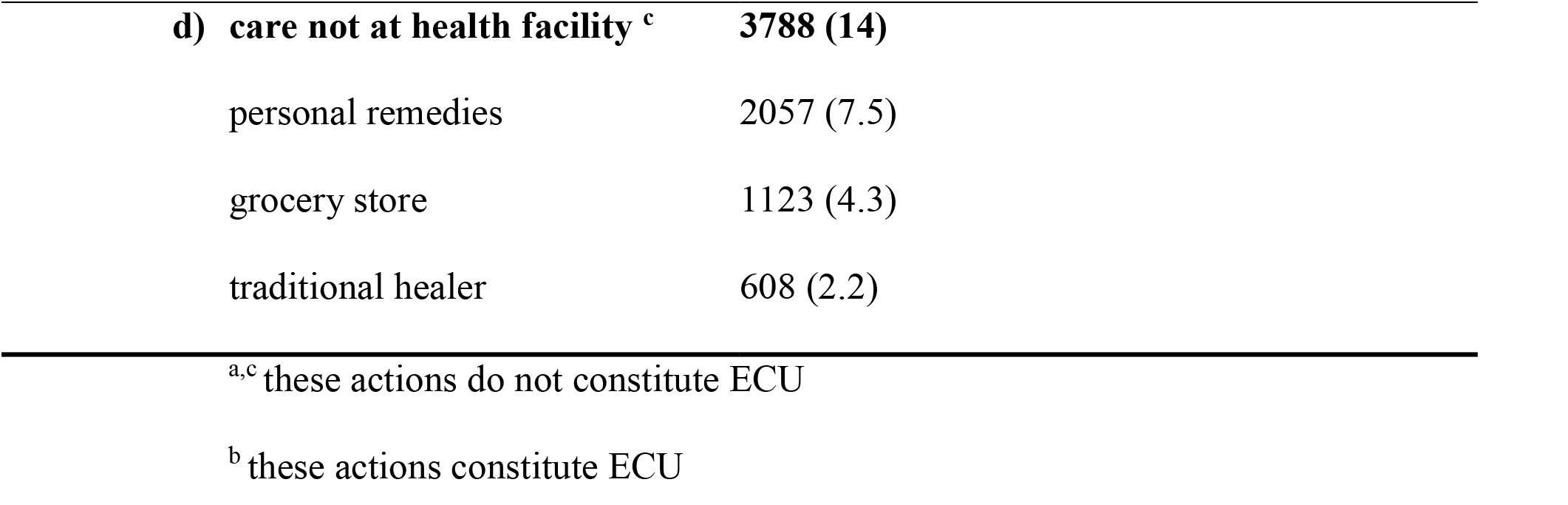
Distribution of actions taken by participants with eye problems

| Action taken (n=27366) | n (%) |
| --- | --- |
| **Did not seek health care <sup>a</sup>** | **6343(25.4)** |
| a) Did nothing, not serious illness | 3393(14.6) |
| b) Did nothing, no money | 2950(10.8) |
| **Sort health care** |  |
| c) **care at health facility <sup>b</sup>** | **16,565 (60.5)** |
| government | 14173(51.8) |
| private | 1316 (4.8) |
| Church/mission | 613(2.2) |
| Pharmacy store | 463 (1.7) |
| **d) care not at health facility <sup>c</sup>** | **3788 (14)** |
| personal remedies | 2057 (7.5) |
| grocery store | 1123 (4.3) |
| traditional healer | 608 (2.2) |

<sup>a,c</sup> these actions do not constitute ECU
<sup>b</sup> these actions constitute ECU

### prevalence of ECU and its associated factors

A total of 16564 (61% of 27366) individuals with eye problems sort eye care. With regards to gender about 5467 (68% of 8014) of male participant’s sort care, compared to 11097 (57 % of 19322) females (p<0.001). According to region, the north had lowest prevalence of ECU 1318 (43 % of 3084) while the south registered the highest 9254 (74% of 12378) (p<0.001). A total of 2733 (66% of 4134) and 13831 (57% of 23202) utilized eye care from the urban and rural areas respectively (p<0.001). And according to level of education, individuals with secondary school education utilized eye services more 4115 (85% of 4828) compared to 7116 (52% of 13577) with no education (p< 0.001). The least frequent age group seeking care at a health facility was older adults (52% of 11155) and the highest was middle aged 4569 (90% of 5063) (p<0.001). According to marital status 10239 (72% of 14140) married people utilized eye care compared to 968 (40 % of 2409) widow/widower. Considering the association between having chronic illness and ECU, 9254 (11.8% of 12378) participants with chronic illness utilized eye care facility compared to 14616 (88 % of 24829) without chronic illness (p<0.001).

A logistic regression was performed to ascertain the effects of residence, region, education qualification, chronic illness, sex and age on the likelihood that participants portray ECU. The model was statistically significant, χ2(8) = 27.402, p <.001. The model explained 34.0% (Nagelkerke R2) of the variance in ECU and correctly classified 73.7% of cases (Table 4). Males were more likely to exhibit ECU than females (OR=1.2,95% CI [1.104,1.290]). But having a chronic condition was associated with a reduction in the likelihood of ECU (OR=0.6, 95% CI [0.547,0.708]). Residing in the urban area was 1.2 times more likely to exhibit ECU than from the rural area (OR=1.2, 95% CI [1.118-1375]). Having a higher education qualification was 6 times likely to seek eye care at a medical facility than not having a formal education (OR=6.6.,95% CI [5.927-7.366]. Middle aged participants were 7 times more likely to utilize eye care services than young adults (OR=7, 95% CI [6.782-8.476]) while older adults were less likely to utilize eye care than middle aged (Table 3).

**Table 3.**
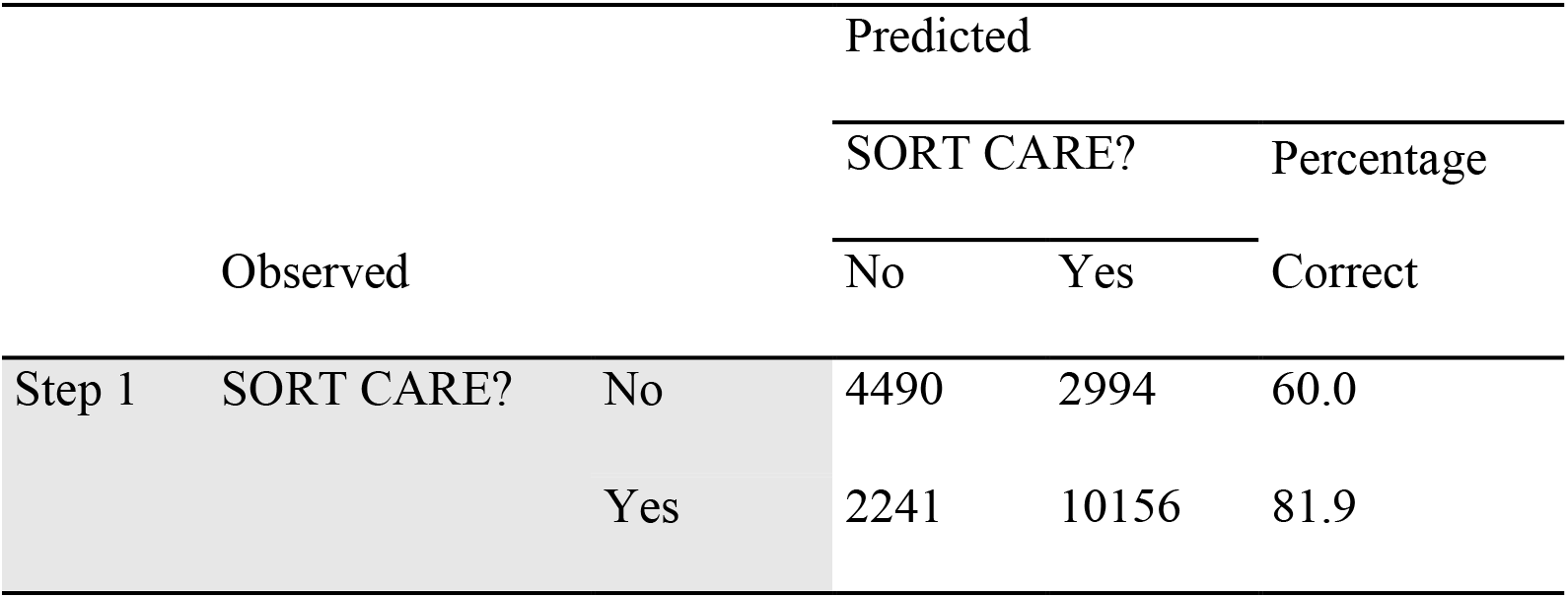
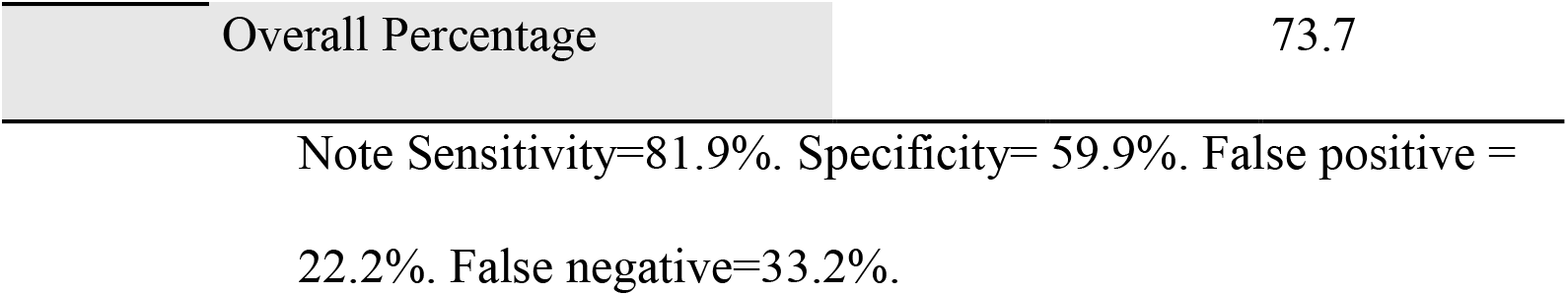
The Observed and the Predicted Frequencies for ECU by Logistic Regression With the Cutoff of 0.50.

| Observed |  |  | Predicted |  |  |
| --- | --- | --- | --- | --- | --- |
|  |  |  | SORT CARE? |  | Percentage |
|  |  |  | No | Yes | Correct |
| Step 1 | SORT CARE? | No | 4490 | 2994 | 60.0 |
|  |  | Yes | 2241 | 10156 | 81.9 |

|  |  |
| --- | --- |
| Overall Percentage | 73.7 |

Note Sensitivity=81.9%. Specificity= 59.9%. False positive = 22.2%. False negative=33.2%.

## Discussion

The rate of ECU varies widely ranging from 18 to 82% as a factor of study settings, and study population. ^[5]^ The prevalence of ECU in our study was similar to a previous report. ^[9]^ However, others reported lower ECU ^[5,14]^ while ECU was higher elsewhere. ^[13,23]^ The rate of ECU in our study could be attributed to the affordability of health services in the country through existing programmes such as EHP.^[15]^ In part, the variation could be due to different operational definitions of ECU. For instance, we defined ECU as seeking care for ocular problem at a medical facility 2 weeks prior to the study, whereas the American Optometric Association (AOA) defines ECU as the use of eye care services in the preceding 3 years. ^[24]^

In our study, the majority of participants who sought eye care services visited government hospitals. On the contrary, others countries reported different main sources of care such as private clinics. ^[5,25]^ The results of our study is not surprising considering that In Malawi, the government is chief actor in health service delivery. ^[15]^

Regarding the reasons for not seeking care, we found that lack of funds is a main hindrance to accessing eye care similar to previous studies. ^[26,27]^ Although in Malawi eye care is provided free of charge, patients still suffer transport costs. ^[28]^

Another larger group of people who did not seek care for their eye problems mentioned negligence as the reason for not seeking care similar to previous research. ^[27]^ This reflects lack of awareness and calls for community sensitization and eye health education programs.

In this review, the rate of self-medication for eye problems was lower than previously reported. ^[18]^. The difference could be due to different sample sizes. Regardless, Self-medication is inherent burden in Africa where majority of eye problems do not go beyond unorthodox alternatives. ^[18]^

Similar to previous report, the rate of ECU was higher among young adults and increased through the middle aged, but dropped among the older adults. ^[29]^ However, it is in disagreement with others. ^[5,30]^ Nevertheless, demand for eye services increases with age since most eye conditions are age related. ^[5]^ Decreased ECU among elderly in our study could be due to lack of an escort to the health facility.^5^

With regards to gender, our study has shown a male predominance similar to previous authors. ^[31.32]^ In contrast, other studies found female predisposition. ^[29,30]^ Nonetheless, Akuwoah and colleagues found no statistically significant difference between gender. ^[26]^ The variation can be explained by cultural differences.

Likewise, previous studies ^[33,34]^ our investigation has also shown that the rate of ECU is higher among those with more education.

Majority of ocular conditions are caused by chronic diseases. ^[10]^ Surprisingly, our study found that people with chronic diseases were less likely to use eye care services, however, others found that chronic diseases are associated with ECU.^[23,35]^ A recent study found that understanding of health and disease is a key determinant of seeking healthcare among persons with chronic conditions in Malawi.^[36]^

Regarding residence, our investigation revealed that people from the urban areas utilize eye care services more than their rural counterparts comparable to previous studies ^[23]^ The rural urban disparity can be explained by less available ophthalmic human resources in rural areas. For instance, in Malawi 11 out of 12 ophthalmologists are based in urban areas. ^[37]^

Concerning region of origin, ECU in Malawi varies with the south registering the highest and north lowest. We cannot explain the regional variation in our study.

A strong caveat of this study is the use of a countrywide population-based data set that provides adequate statistical power to reconnoiter the association between ECU and its associated factors. Nevertheless, our study is not without drawbacks. First, the study is based on subjective responses which are prone to recall bias by the study participants. Another important caveat of our study is that we did not include the association between economic status and ECU including wealth and OPE.

## Conclusion

In summary, we discussed a novel report of ECU and its associated factors among a population of Malawian adults using nationally representative data. The results demonstrate that in Malawi access to eye care services is entrenched in social disadvantages. Specifically, ECU Is low among older adults. Rural areas, women, and the less educated. We recommend strategies to improve social support, Unequal distribution of resources between urban and rural areas, education opportunities for all, women empowerment, and task shifting approach. Further investigations can elucidate ECU usage among children in Malawi.

## Data Availability

All data produced are available online at https://microdata.worldbank.org/index.php/catalog/2939.

